# Improving dengue case confirmation by combining rapid diagnostic test, clinical, and laboratory variables

**DOI:** 10.1101/2021.05.21.21257609

**Authors:** Carolina Coronel-Ruiz, Myriam L. Velandia-Romero, Eliana Calvo, Sigrid Camacho-Ortega, Shirly Parra-Alvarez, Edgar Beltrán-Zuñiga, María-Angélica Calderón-Pelaez, Fabián Cortés-Muñoz, Juan Pablo Rojas-Hernandez, Syrley Velasco-Alvarez, Alfredo Pinzón-Junca, Jaime E. Castellanos

**Affiliations:** Universidad El Bosque, Vicerrectoría de Investigaciones. Grupo de Virología. Bogotá, Colombia; Universidad El Bosque, Vicerrectoría de Investigaciones. Bogotá, Colombia; Fundación Clínica Infantil Club Noel. Cali, Colombia; Hospital Universitario de La Samaritana. Bogotá, Colombia; Hospital Universitario de La Samaritana. Grupo Investigación en Riesgo Cardio-Vascular, Trombosis y Anticoagulación (RICAVTA). Bogotá, Colombia

## Abstract

**Background:** Dengue is the most widely distributed arboviral disease in tropical and subtropical countries. Early diagnosis is difficult, and most of the suspected cases are diagnosed according to clinical criteria. In underdeveloped countries, laboratory tests are done on a proportion of dengue with warning signs or severe dengue suspect cases. This study aimed to design a diagnostic algorithm using rapid diagnostic tests (RDT), ELISA tests together with clinical and hematology variables to confirm dengue cases in febrile patients from an endemic area in Colombia.

**Methods and results:** A total of 505 samples were collected from patients with acute febrile syndrome (<*7* days) assisted to the Municipal Hospital in Girardot (Colombia). Serum samples were evaluated by rapid diagnostic tests -RDT- (IgM and IgG antibodies and NS1 antigen immunochromatographic assay), capture ELISAs (IgM, IgG, and NS1 antigen), and by RT-PCR. We analyzed individual tests performance to determine which were the most useful to confirm dengue cases. Individual results for IgM, IgG, and NS1 RDT yield low sensitivity and specificity values than the reference standard. A high sensitivity (96.3%) and specificity (96.4%) were obtained after combining the IgM and NS1 ELISAs results. The analysis using the combined NS1 RDT and IgM ELISA results showed 90.3% sensitivity and 96.2% specificity. Adjusted odd ratios (aOR) were calculated including data from symptoms, signs, and diagnostic laboratory tests to differentiate dengue from other febrile illnesses (OFI). Myalgia (aOR: 1.87, CI95%: 1.04-3.38), abdominal tenderness (aOR: 1.89, CI95%: 1.14-3.10), platelets count <140.000/mm^3^ (aOR: 2.19 CI95%: 1.31-3.67). The analysis using the results of the diagnostic test yields significant ratios for IgM RDT (aOR:2.63 CI95%: 1.59-4.33) and NS1 RDT also differentiate dengue cases from OFI. Combined positive IgM or NS1 RDT and the one that combined positive NS1 RDT or IgM ELISA detect dengue cases in 81.6% and 90.6%, respectively (p<0.001).

**Conclusion:** Our findings showed that only clinical diagnosis does not confirm true dengue cases dengue and needs to be complemented by laboratory diagnostic tests to stablish the diagnosis. We also demonstrate the usefulness of rapid tests in diagnosis, suggesting their implementation with IgM ELISA test to better confirm dengue cases.

**Author Summary:** Dengue infections are considered the main mosquito-borne viral disease and a considerable concern in public health in tropical countries. Transmission has been associated with population growth, globalization (rapid urbanization, increasing movement of people), and environmental changes. Dengue diagnosis is difficult because of clinical variability in population of different age groups, with different degrees of severity that can have a fatal outcome. Diagnosis must consider clinical and laboratory criteria to improve early case confirmation particularly in endemic areas, where there is a need to support health services and improve treatment and management of the disease.

## Introduction

Dengue is the most important human arbovirosis in tropical and subtropical countries. It is caused by one of the four different dengue virus (DENV) serotypes (DENV-1 to DENV-4) and is transmitted after the bite of *Aedes* mosquitoes circulating in Asia and Latin-American countries [1-5]. During the initial stage, patients with dengue show many unspecific signs and symptoms, similar to other infectious diseases such as influenza, chikungunya fever, zika fever, leptospirosis, or yellow fever [6-10]. As the disease progresses, it can follow variable clinical courses, ranging from mild fever to severe and complicated symptoms, which may have a fatal outcome [11-14].

Early diagnosis of dengue infection is difficult, most of the infections are not identified or are delayed, leading to complications in patients [1, 15, 16]. In the last decade, dengue diagnosis is made following the World Health Organization (WHO) definition of dengue cases proposed in 2009 [17, 18] many health care centers and international organizations have been involved in studies to establish dengue diagnostic protocols integrating both clinical criteria and laboratory tests [19, 20]. Simultaneously, several investigations have been carried out to design diagnostic algorithms, which aim to improve the case confirmation and strengthen overall disease management [21-26]. In underdeveloped countries, in addition to inequality in the access to health services, the inability to perform diagnostic tests on the same day of sample collection due to lack of equipment or trained personal or high costs remains the main drawback of the dengue surveillance system [4, 27, 28].

Colombia has a high number of dengue cases and has the second-highest prevalence rate in the Americas [29-31] after Brazil [32-34]. This is due to *Aedes aegypti* infestation in urban areas and its wide circulation in more than 70% of the municipalities located in areas below 1.800 MASL. Recently, *Aedes* mosquito has been identified in rural zones [29, 35, 36] and also in some municipalities in areas above 2200 MASL. In addition, steady circulation of all four dengue serotypes has been confirmed in many municipalities [31, 37]. This has led to dramatic changes in the dengue endemicity with dengue cases being reported in municipalities with no prior cases, reduction in the mean age of patients suffering from dengue [29, 38], and an increasing number of cases with severe dengue and fatalities in the population under 15 years of age [39, 40]. This pattern is similar to that observed in countries like Nicaragua [41, 42] and Southeast Asian countries like Vietnam and Thailand, where dengue is highly prevalent [43]. Together, these different factors may not only help in understanding the rising incidence and prevalence rates in our country but also explain the increasingly frequent severe and fatal cases [30] Unfortunately, most of the reported dengue cases in Colombia are diagnosed by clinical criteria, and only a small percentage of the cases are confirmed by Immunoglobulin M (IgM) enzyme-linked immunosorbent assay (ELISA) test or reverse transcription-polymerase chain reaction (RT-PCR) (34.7%); only severe or fatal cases are investigated by non-structural protein-1 (NS1) ELISA or RT-PCR [44, 45]. For these reasons, some secondary and tertiary care hospitals independently perform rapid diagnostic tests (RDT) for IgM/IgG or NS1 to improve the clinical diagnosis and treatment of cases suspected of dengue [46-48]. Despite the widespread use of RDT in Colombia, the few reports evaluating the performance of RDT suggest that a negative result in these tests does not rule out dengue; therefore, clinicians should be aware of dengue diagnosis and confirm cases by laboratory tests [47-50].

Due to the shortcomings in implementing laboratory tests in underdeveloped countries, in the last few years, many studies in Asia and the Americas have focused on the evaluation of clinical signs or symptoms and assigning scores depending on the frequency or severity to establish the final diagnosis [12, 21-23, 51]. Although this strategy has been helpful in the early identification of cases and improving patient care and treatment, the validation of these clinical algorithms is difficult due to the heterogeneity of the study population, the variability in the clinical presentation between countries, and the differences in the signs or symptoms according to the disease stages [52-59]. Other studies have combined clinical variables with hematological or biochemical data for confirmation of cases, such as those reported in Colombia and Vietnam [24, 60]; a predictive model proposed in Brazil [52] found redness of eyes and leukocyte count as key variables to differentiate dengue from other febrile illnesses.

Most reported studies have evaluated the performance of serological or virology tests against one or more tests while integrating a gold standard diagnostic test. These studies show high variability in the sensitivity, specificity, and predictive values due to factors like patient population enrolled, differences in the DENV serotypes, timing of sample collection, and the test or tests considered as reference standards. However, these studies shed light on the usefulness of these approaches for dengue case confirmation and establish their potential as predictors of the disease severity [34, 61-69].

As an endemic country, Colombia requires new strategies to strengthen the clinical diagnosis and the laboratory confirmation for dengue cases and evaluate the sensitivity. and specificity of tests that can be used in every health care center independently of their complexity. This may improve the early detection of cases and more efficiently control the outbreaks, while simultaneously strengthening the dengue surveillance system. Therefore, this study aimed to design a diagnostic algorithm using RDT and clinical and hematological variables to identify and confirm dengue cases in febrile patients from an endemic area in Colombia.

## Methods

### Patients and data collection

This study was revised and approved by the Ethical Committee of Hospital Universitario de La Samaritana, Bogotá (Resolution 7 and 9 of 2013 and 2014 respectively). Patients consulting for a primary diagnosis of febrile syndrome in the Hospital de Girardot from March 2014 to July 2015 were enrolled. The Girardot municipality is located 134 km from Bogotá, has 150 000 inhabitants, is located at an altitude of 289 MASL with a mean temperature of 33.3°C, and 66.4% of relative humidity. Girardot reported a dengue incidence of 572.5 cases [95% confidence interval, (CI 525.3 – 619.9)] and 98 000 individuals at risk in 2010 [70].

Patients were invited to participate and signed informed consent or an informed assent form (in case of children with a consenting guardian). The inclusion criteria were patients with fever onset less than 7 days, malaise, headache, retro-orbital pain, myalgias, exanthema, abdominal pain, and arthralgias, following the disease description by WHO in 2009 [17]. Patients with an apparent infectious focus were excluded (otitis, tonsillitis, or urinary infection). Sample size was calculated based on acceptable precision for estimates of sensitivity and specificity values and the estimated prevalence of disease in a endemic area.

### Sample processing

After a complete medical examination and completing the clinical report form, blood samples were taken for immediate analysis of hematological variables (leukocyte, erythrocytes, platelet counts, hemoglobin, and hematocrit) and the sera were processed for evaluation by RDT for IgM/IgG antibodies [Dengue duo cassette (01PF10), Panbio, Alere] and NS1 RDT [Dengue early rapid (01PF20), Panbio Alere]. The remaining serum was aliquoted, frozen, sent to the Laboratory (Universidad El Bosque, Bogotá), and stored at -80 °C until further use. All the samples were processed for RNA extraction (QIAmp viral mini-kit, Qiagen) following the manufacturer’s instructions. The isolated RNA was retrotranscribed and amplified using primers described previously [71] and the PCR modified protocols [72]. For the first round, SuperScript III Platinum One-step RT-PCR (Invitrogen) and, mD1 and D2 primers were used; the amplified product was analyzed for the presence of DENV serotypes in a multiplex format using specific primers mD1, TS1, mTS2, TS3, and rTS4. The final reaction products were separated in 2% agarose gel, stained with ethidium bromide, and analyzed.

IgM Capture ELISA [UMELISA Dengue IgM Plus (UM2016), Tecnosuma Intl, La Havana] and IgG Capture ELISA [Panbio Dengue IgG Capture ELISA (01PE10)] were used to confirm the cases; positive results for either of these tests were further analyzed by NS1 antigen ELISA [Panbio Dengue Early ELISA (01PE40)]. Additionally, indirect IgG ELISA [Panbio Dengue IgG Indirect ELISA (01PE30)] was performed to assess the history of dengue infection in enrolled patients. Around 10% of the patient provided a second serum sample during convalescence (10-30 days after acute phase), and these samples were processed for IgM and IgG Capture ELISA to determine seroconversion.

### Clinical diagnosis and dengue case confirmation

Dengue cases were clinically diagnosed and classified following the WHO 2009 criteria [17]. Additionally, hematological parameters were also assessed in the following groups: dengue, dengue with warning signs (DWS), and severe dengue (SD); however, the final classification was based on the laboratory tests. A confirmed dengue case was one that was positive by IgM Capture ELISA or RT-PCR or if there was a seroconversion between the acute and convalescent samples. The remaining samples were categorized as Other Febrile Illnesses (OFI). Additionally, positive samples in the IgG Capture ELISA tests were classified as secondary infections. Finally, to establish the percentage of true dengue cases, an algorithm integrating the clinical description with laboratory findings was derived and was divided into three different diagnostic categories: dengue, DWS, and SD.

### Statistical analyses

Quantitative variables measuring central tendency (averages) and dispersion (standard deviation) were employed, after checking the normality of their distribution by means of a Shapiro–Wilk test. If the assumption was not verified, they were described using mean and interquartile ranges. Furthermore, qualitative variables were described using proportions. To compare the difference between groups (i.e., according to diagnosis), a one-way ANOVA was utilized for continuous variables with normal distribution, or failing this, nonparametric statistics were used (e.g., Kruskal - Wallis test). In the case of qualitative variables, differences were calculated by Pearson’s chi-square test when the expected square values were ≥5. Otherwise, Fisher’s exact test was applied. Dengue diagnostic test sensitivity, specificity, predictive values were determined, receiver operating characteristics (ROC) curves were derived, and area under curve (AUC) was estimated to compare the dengue laboratory tests and the confirmation case algorithm or if there was an improvement after combining a few parameters from them.

Odds ratios (OR) were calculated to identify the confounding clinical variables in the final diagnosis and to identify useful valuable variables to build a logistic regression model to measure the strength of association between signs and symptoms and the final diagnosis outcome: dengue, DWS, and SD. A regression analysis was performed to calculate the adjusted odds ratios (aOR) and 95% CI; models presenting p< 0.05 were included to propose the decision trees. Model’s reliability was evaluated through re-estimation tests, the Deviance test, and the Hosmer–Lemeshov goodness of fit test. A *p* value <0.05 was considered statistically significant. Statistical analyses were performed using Stata V.12.1.

## Results

Clinical and laboratory data on 505 patients were evaluated (54.7% male); the median age was 11 years [Interquartile range (IR 6 - 23)], with 80.6% of the patients presenting within the first seven days of the illness. Only 21.9% had a temperature above 38°C during the first consultation (Table 1). Following the clinical criteria suggested by WHO and the results of defined laboratory dengue tests (ELISA IgM or RT-PCR or seroconversion), a definitive diagnosis was assigned to each patient; there were 305 (76.2%) confirmed cases of dengue (19% Dengue, 45.7% DWS, and 11.5% SD cases). Patients negative for the selected tests were classified as OFI (23.8%).

**Table 1.**
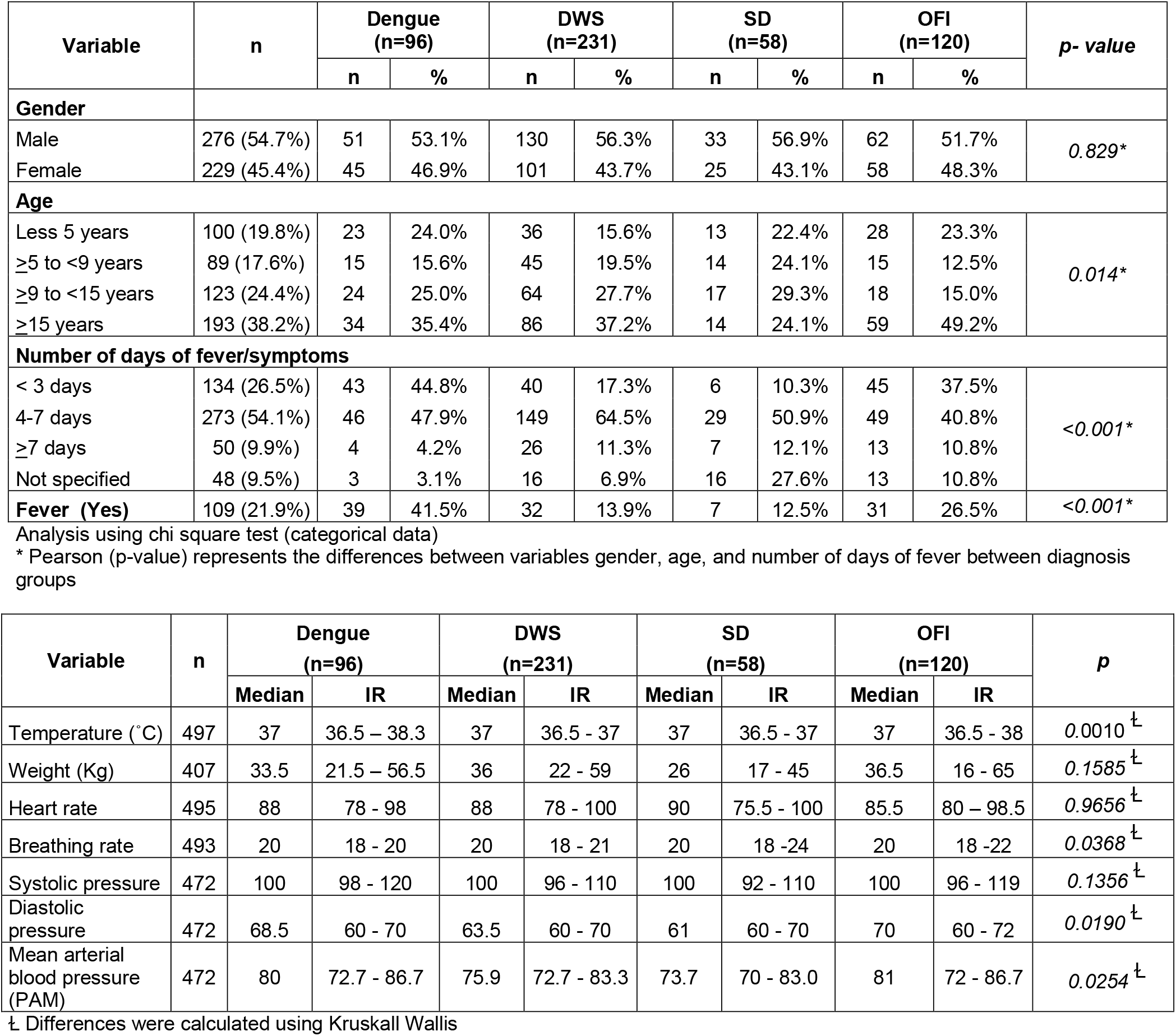
Demographic characteristics, signs and symptoms of dengue cases and OFI identified

The most frequent symptoms reported were myalgia (81.8%), headache (73.5%), abdominal pain (61.8%), nausea (53.8%), vomiting (53.6%), and retro ocular pain (31%). The main identifiable signs were painful abdomen on palpation (54.6%), hepatomegaly (14.1%), ascites (10.9%), and edema (10.3%). The leukocyte count was <4 000/mm^3^ in 34.9% of the patients while platelet counts <100 000/mm^3^ were observed in 43.7% of the patients (Table 2). With the dengue RDT, 64.1% and 66.1% of the samples were positive for dengue IgM and IgG, respectively; however, only 30.5% of the samples were positive for Dengue Early Rapid test NS1 RDT. The IgM Capture, IgG Capture, and NS1 ELISAs were positive in 63.4%, 52.4%, and 30.5% of the patients, respectively. Around 86% of the patients showed a previous contact with dengue (IgG indirect positive) (Fig 2).

**Table 2.** Laboratory values according to diagnosis.

| Variable | Dengue<br>(n=96) | DWS<br>(n=231) | SD<br>(n=58) | OFI<br>(n=120) | p |
| --- | --- | --- | --- | --- | --- |
| Leukopenia<sup>†</sup><br>(Yes)<br>176 (34.9%) | 42 (43.8%) | 83 (36.1%) | 16 (27.6%) | 35 (29.2%) | 0.089 |
| WBC Count<br>($\times 10^3/\text{mm}^3$) | 4.3 (3.5 - 5.8) | 4.9 (3.4 - 7.1) | 5.9 (3.8 - 8.5) | 5.1 (3.8 - 7.5) | 0.0178<sup>‡</sup> |
| Thrombocytopenia<sup>†</sup><br>(Yes)<br>220 (43.7%) | 20 (20.8%) | 136 (59.1%) | 37 (63.8%) | 27 (22.5%) | <0.001 |
| Platelets<br>($\times 10^3/\text{mm}^3$) | 135.5 (107 - 174) | 88 (63 - 121.8) | 83.2 (52 - 115) | 140 (102.3 - 194) | 0.0001<sup>‡</sup> |
| RBC Count<br>($\times 10^3/\text{mm}^3$) | 4.6 (4.3 - 4.8) | 4.6 (4.3 - 4.9) | 4.6 (4.3 - 5.2) | 4.4 (4.0 - 4.7) | 0.0002<sup>‡</sup> |
| Haemoglobin<sup>†</sup><br>(g/dL) | 12.8 (11.9 - 13.7) | 12.7 (11.8 - 14.2) | 12.7 (11.8 - 14.2) | 12.2 (11.3 - 13.3) | 0.0042<sup>‡</sup> |
| Haematocrit<sup>‡</sup> (%) | 38.2 ± 3.9 | 38.3 ± 5.8 | 37.3 ± 5.8 | 36.5 ± 5.0 | 0.005<sup>†</sup> |
| Neutrophils (%) | 49.7 (30.4 - 63.7) | 37.0 (25.1 - 55.2) | 40.2 (28.0 - 55.9) | 57.0 (45.7 - 70.3) | 0.0001<sup>‡</sup> |
| Lymphocytes (%) | 39.5 (24.8 - 57.2) | 51.1 (34.2 - 63.7) | 50.8 (35.1 - 61.4) | 32.6 (18.7 - 43.2) | 0.0001<sup>‡</sup> |
| Monocytes (%) | 8.4 (6.6 - 9.8) | 8 (5.6 - 10) | 6.9 (4.9 - 9.1) | 7.9 (5.9 - 9.8) | 0.0465<sup>‡</sup> |
| Eosinophils (%) | 0.6 (0.2 - 2.8) | 1.3 (0.4 - 3.0) | 0.7 (0.4 - 1.7) | 1.0 (0.3 - 2.2) | 0.0848<sup>‡</sup> |
| Basophils (%) | 0.4 (0.3 - 0.7) | 0.6 (0.3 - 1.1) | 0.5 (0.2 - 0.9) | 0.3 (0.2 - 0.4) | 0.0001<sup>‡</sup> |

Data are presented median (interquartile range), <sup>†</sup>n (%) values, <sup>‡</sup> media ± SD.
Differences were calculated using <sup>‡</sup> Kruskal Wallis <sup>†</sup> One way ANOVA

**Figure 1.**
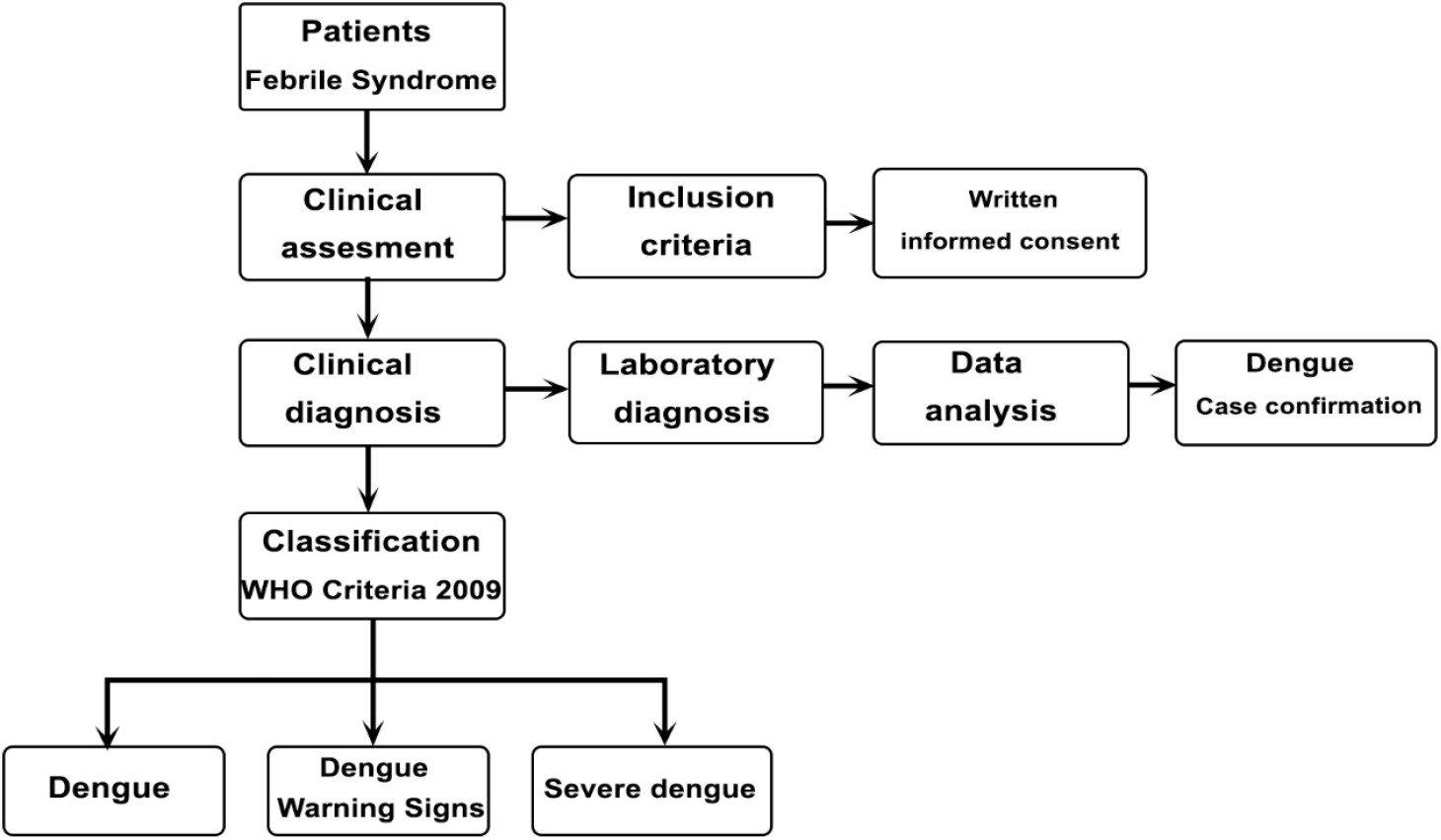
Flowchart dengue clinical diagnosis, case confirmation, and classification according to severity of the disease

**Fig 2.**
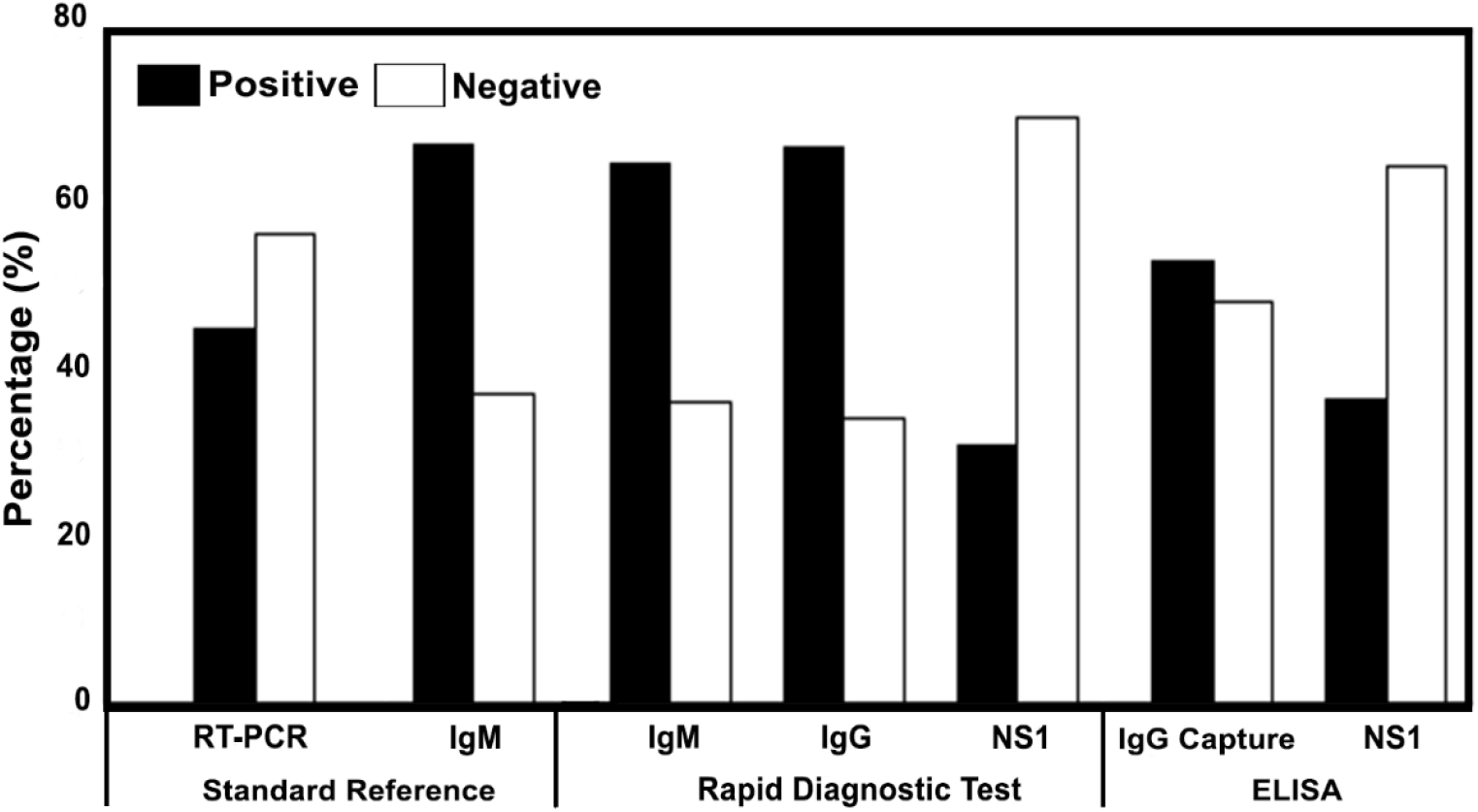
Frequency of positive and negative samples analyzed. Samples from febrile phase were processed to identify dengue cases. Rapid Diagnostic Tests (RDT) for IgM and IgG antibodies, and NS1 antigen were applied in health institution and their results support clinical diagnosis. Samples were evaluated with ELISA tests and RT-PCR to stablish laboratory diagnosis. IgM ELISA and RT-PCR were considered as standard reference

Clinical manifestations such as myalgia (84.2% vs 73.8%), rash (27.1% vs 8.8%), and abdominal pain (55.7% vs 45.9%) were significantly more frequent in dengue confirmed cases than the OFI group, respectively (p<0.05). For example, 43.2% of patients in the OFI group had abdominal tenderness on palpation compared to 74.8% in DWS and 80.4% in the SD group. Other symptoms like hepatomegaly and ascites were significantly more frequent in dengue cases than OFI patients. For example, hepatomegaly was identified in 18.5% patients in the DWS group and 42.1% in the SD group vs 3.4% in the OFI group *(p<0*.*001);* edema was seen in 11.7% of the patients in the DWS group and 29.8% in the SD group vs 6.7% in the OFI group *(p<0*.*001)*. On the other hand, exanthema were reported in fewer patients in the entire cohort (10.1%, 5.2%, 3.6%, and 2.5% in DWS, Dengue, SD, and OFI groups, respectively) (Fig 3). However, the OFI group in the present study cohort showed respiratory difficulty (5.0%) and pleural effusion (2.5%), two characteristic clinical signs commonly seen in patients with severe dengue (72.4% and 100%, respectively). Platelet counts were significantly lower in the dengue confirmed cases compared to the OFI group *(p=0*.*0001)*.

**Fig 3.**
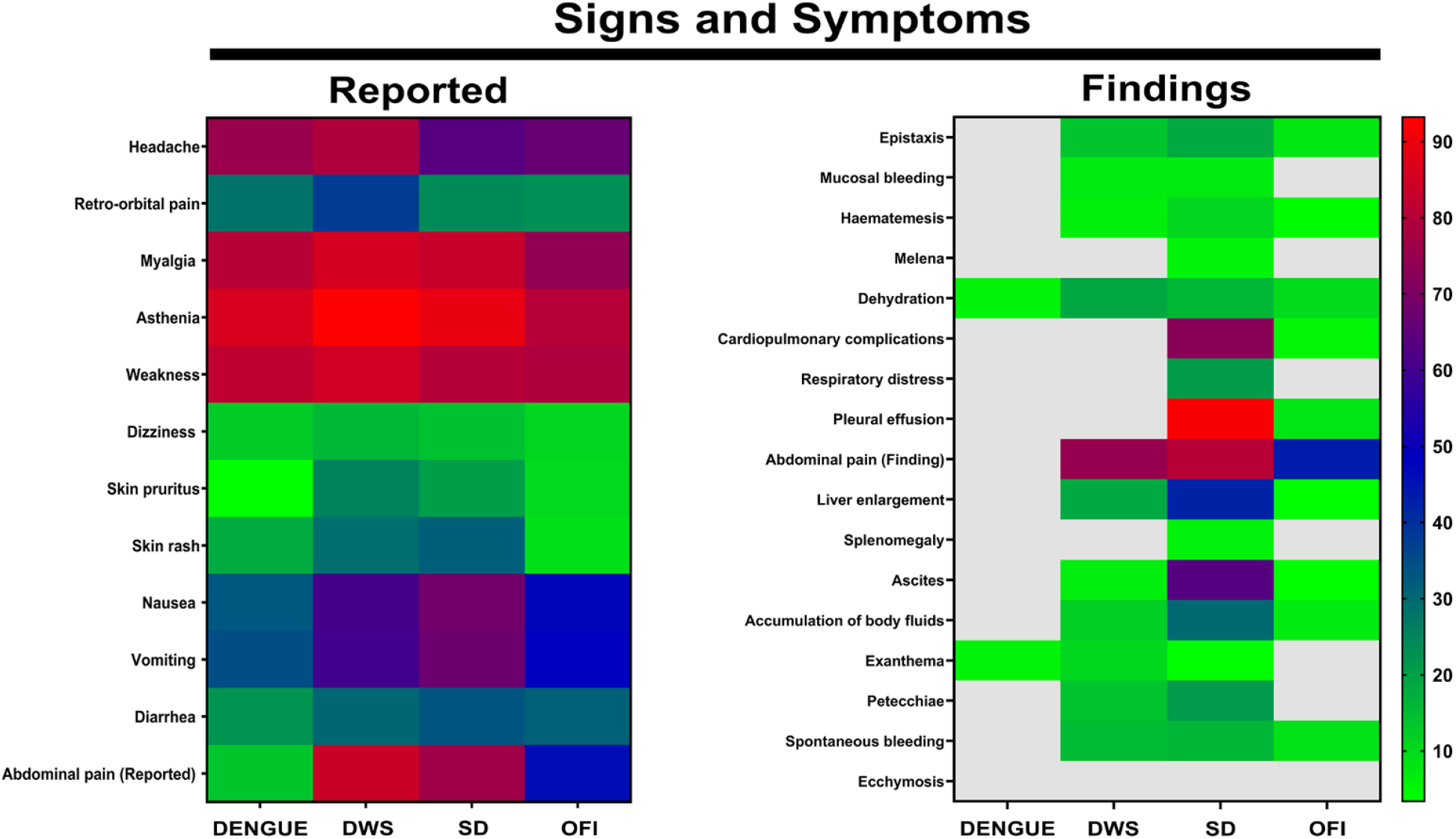
Clinical features of febrile patients according to diagnosis and disease severity. Maps present clinical information from febrile patients included in the study, according to diagnosis: Dengue without warning signs (DENGUE), dengue warning signs (DWS), severe dengue (SD), Other Febrile Illness (OFI). Colors represent the percentage of patients presenting specific clinical sign.

### Performance of dengue laboratory tests

Considering true dengue cases as the reference, we compared the performance of the RDT, and other tests performed in the study. Positive IgM RDT was significantly lower in Dengue patients (60%) compared to DWS (73.2%) or SD cases (77.2%) (p=0.029); similar results were observed for IgG RDT (56.8% of dengue samples were positive while 77.1% and 80.7% were positive in the DWS and SD groups, respectively; *p<0*.*0001)*. However, there was no difference in the number of positive patients between the groups with NS1 RDT *(p=0*.*612)*. The percentage of IgM Capture ELISA positive cases was significantly different between the groups (64.1%, 86.8%, and 89.1% for dengue, DWS, and SD, respectively; *p<0*.*001*). Similar results were observed for the IgG Capture ELISA where 39.1%, 68.1%, and 58.9% cases were positive in the dengue group, DWS, and SD, respectively (p<0.001). The NS1 ELISA test results did not show differences between the groups *(p=0*.*630)*. On the other hand, RT-PCR detected significantly more dengue cases (70.2%) than DWS (55.2%) or SD cases (50.0%) *(p=0*.*018)*. The RT-PCR results indicated that DENV-2 was the most frequently identified serotype in the dengue, DWS, and SD groups (69.7%, 63.0%, and 58.6%, respectively), followed by DENV-1 (12.1%, 14.2%, and 17.2% in dengue, DWS, and SD group, respectively). We also observed that DENV-1/DENV-2, DENV-1/DENV-3, and DENV-2/DENV-3 coinfections were slightly higher in the SD cases (13.8%) than DWS and the dengue group (13.4% and 9.1%, respectively) (S1 Fig). Based on indirect IgG ELISA, most of the cases were secondary infections in the three groups: dengue, DWS, and SD groups (85.4%, 90.9%, and 89.5% respectively) *(p=0*.*342)*.

### Quantitative analysis of dengue diagnostic tests

We analyzed different sets of tests to determine the one that was most accurate to confirm dengue cases. The sensitivity, specificity, predictive values, and ROC area under the curve (AUC) were estimated for IgM, IgG, NS1 RDT, IgG Capture, and NS1 ELISA using confirmed dengue cases as a reference standard (positive samples for IgM Capture ELISA or RT-PCR); the individual sensitivity and specificity were 81.6% for capture IgM ELISA and 58.1% for RT-PCR.

Individual IgM, IgG, and NS1 RDT results did not yield superior values than the reference standard; similar results were seen after the analysis with IgG capture and NS1 ELISAs (S1 Table). However, the sensitivity (97.0 %), specificity (82.1%), and ROC AUC (0.896) after combining IgM Capture and NS1 ELISAs were higher than the values obtained with individual tests. However, these results were not replicated when IgM Capture ELISA was combined with IgG Capture ELISA, or when NS1 ELISA was combined with IgG Capture ELISA during the analysis. To complement this information, combining the results of NS1 RDT and capture IgM ELISA showed better values (90.3%, 96.2%, and 0.932 for sensitivity, specificity, and AUC, respectively) than those obtained for each one of the tests that constituted the reference standard (Fig 4). Further, the positive results from RDT (IgM or IgG or NS1) combined with the RT-PCR results showed a higher sensitivity (97.4%) but low specificity (36.3%) and AUC (0.668).

**Figure 4.**
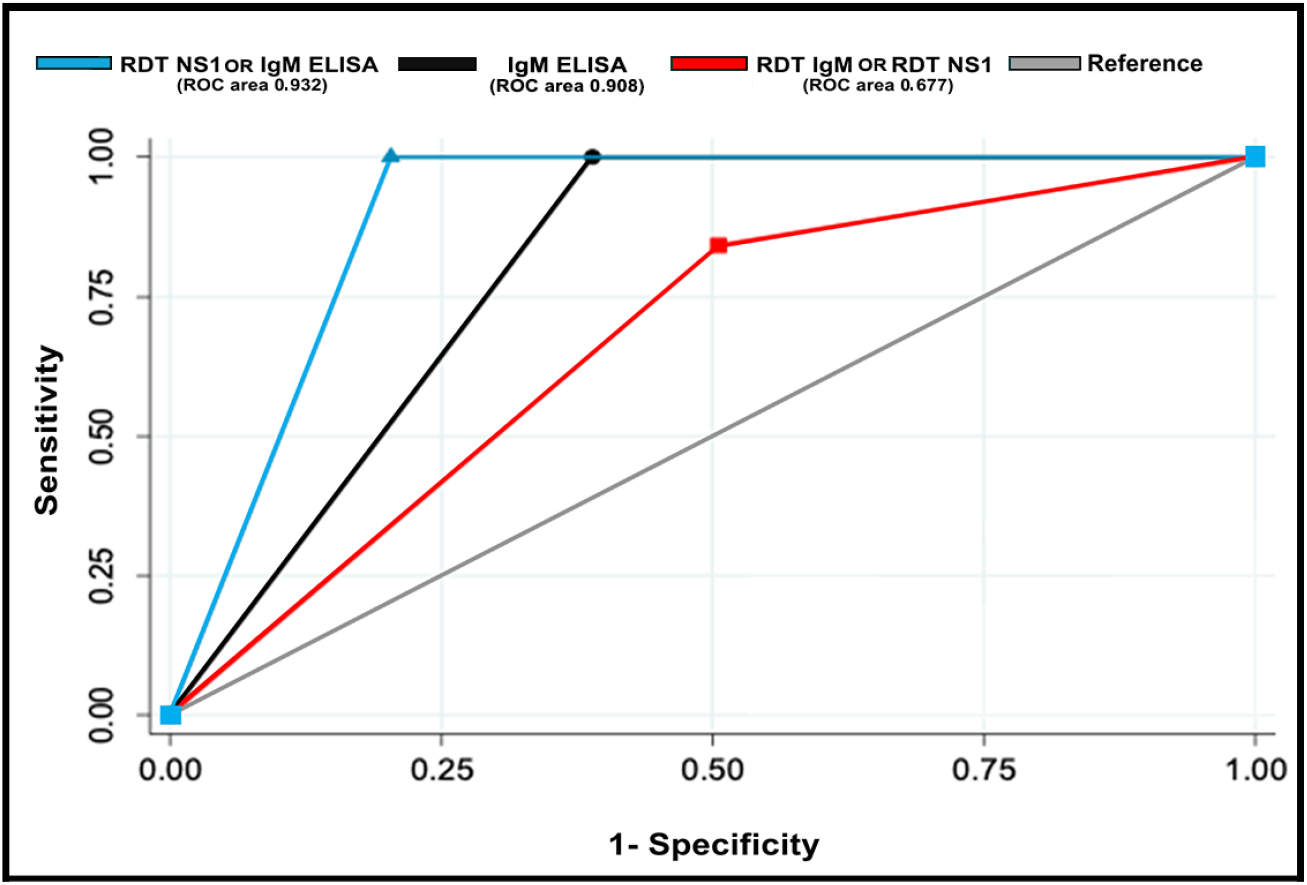
Receiver operating characteristic (ROC) curves of the combination of the dengue diagnostic tests compared to standard reference.

On the other hand, analysis using a combination of any of the positive RDT (IgM OR IgG OR NS1) showed a higher sensitivity (90.5%) but lower specificity (37.4%) compared to ELISA tests. Also, when we evaluated the combination of results of two RDT with one of them being positive (IgM OR IgG; IgM OR NS1; IgG OR NS1), the sensitivity and specificity identified were 69.2% and 71.3%, respectively. However, we observed a significant decrease in the sensitivity (22.1%) and an increase in the specificity (99.1%) by combining positive test results of the three RDT applied (S1 Table). We did not find improvement in the sensitivity or specificity in confirming dengue cases upon evaluating a combination of leukocytes or platelets counts with the results of individual dengue diagnostic tests, compared to the reference standard.

### Dengue cases confirmation predicting models

Using the median and interquartile ranges of leukocytes and platelets of dengue confirmed cases and OFI patients, we established a dengue-related disease cutoff of 5100 cells/mm^3^ and 140000 platelets/mm^3^. This data was used to perform a univariate analysis and calculate the odds ratio (OR) for symptoms, signs, and laboratory diagnostic test results. This approach showed that myalgia, abdominal tenderness, platelets <140000/mm^3^, and the results of the RDT (alone or in combination) were identified with the higher sensitivity to confirm dengue cases (Table 3). On the contrary, for this Colombian cohort, clinical signs such as fever, exanthema, leukopenia <5100/mm^3^, days of disease evolution, and age were not significantly associated with the dengue diagnosis. After the univariate analysis, variables with p-values less than 0.05 were included in the logistic regression analysis to determine the effect of these variables on the diagnosis of dengue. The adjusted OR estimates (aOR) showed that myalgia (aOR: 1.87, 95% CI: 1.04-3.38, *p=0*.*038*), abdominal tenderness (aOR: 1.89, 95% CI: 1.14-3.10, *p=0*.*013*), platelets count less than 140 000/mm^3^ (aOR: 2.19, 95% CI: 1.31-3.67 *p=0*.*003*), and IgM RDT positive result (aOR: 2.63 95% CI: 1.59-4.33, *p<0*.*001*) were independent variables enabling a differential dengue diagnosis compared to OFI. The positive NS1 RDT results adjusted for these signs also enabled in differentiating dengue cases from OFI cases (Table 4).

**Table 3.** Univariate analysis of clinical and laboratory variables as possible predictors of dengue diagnosis.

| Variable | Univariate analysis |  |  |  |  |  |  |  |  |  |  |  |
| --- | --- | --- | --- | --- | --- | --- | --- | --- | --- | --- | --- | --- |
|  | Dengue cases vs.OFI |  |  | Dengue vs. OFI |  |  | DWS vs. OFI |  |  | SD vs.OFI |  |  |
|  | OR | CI 95% | p | OR | CI 95% | p | OR | CI 95% | p | OR | CI 95% | p |
| Fever (Yes) | 0.72 | 0.433 - 1.20 | 0.1724 | 1.97 | 1.06 - 3.67 | 0.0215 | 0.45 | 0.25 - 0.81 | 0.0040 | 0.39 | 0.14 - 1.012 | 0.0375 |
| Myalgia (Yes) | 1.89 | 1.08 - 3.26 | 0.0152 | 1.47 | 0.69 - 3.22 | 0.2903 | 2.14 | 1.15 - 3.95 | <0.001 | 1.73 | 0.71 - 4.55 | 0.1946 |
| Abdominal pain Reported (Yes) | 2.36 | 1.49 - 3.74 | 0.0001 | 0.18 | 0.08 - 0.39 | <0.001 | 6.13 | 3.53 - 10.65 | <0.001 | 3.81 | 1.76 - 8.58 | 0.0002 |
| Abdominal pain Finding (Yes) | 1.84 | 1.17 - 2.89 | 0.0052 | 0 | 0 - 0.06 | <0.001 | 3.89 | 2.34 - 6.48 | <0.001 | 5.37 | 2.39 - 12.64 | <0.001 |
| Orbital pain (Yes) | 1.68 | 0.95 - 3.05 | 0.0619 | 1.29 | 0.57 - 2.88 | 0.5036 | 2.02 | 1.10 - 3.77 | 0.0152 | 1.05 | 0.41 - 2.58 | 0.9130 |
| Edema (Yes) | 1.79 | 0.80 - 4.55 | 0.1382 | 0 | 0 - 0.56 | 0.009 | 1.84 | 0.78 - 4.83 | 0.1424 | 5.89 | 2.18 - 16.89 | <0.001 |
| Exantema | 3.30 | 0.99 - 17.20 | 0.0401 | 2.12 | 0.39 - 13.99 | 0.3001 | 4 | 1.27 - 22.97 | 0.0110 | 1.41 | 0.11 - 12.62 | 0.7121 |
| Platelets <140.000/mm<sup>3</sup> | 3.02 | 1.93 - 4.72 | <0.001 | 1.17 | 0.66 - 2.08 | 0.5631 | 4.63 | 2.75 - 7.78 | <0.001 | 4.42 | 2.002 - 10.302 | <0.001 |
| Leukocytes <5.100/mm<sup>3</sup> | 1.29 | 0.84 - 1.98 | 0.2267 | 2.27 | 1.25 - 4.15 | 0.0038 | 1.17 | 0.73 - 1.86 | 0.4909 | 0.78 | 0.39 - 1.54 | 0.4476 |
| **Diagnostic tests** |  |  |  |  |  |  |  |  |  |  |  |  |
| RDT |  |  |  |  |  |  |  |  |  |  |  |  |
| IgM | 3.15 | 2.01 - 4.95 | <0.001 | 1.98 | 1.10 - 3.57 | 0.0146 | 3.59 | 2.19 - 5.91 | 0.0146 | 4.47 | 2.08 - 9.97 | <0.001 |
| IgG | 3.26 | 2.07 - 5.12 | <0.001 | 1.62 | 0.91 - 2.91 | 0.0825 | 4.133 | 2.49 - 6.85 | 0.0000 | 5.15 | 2.32 - 12.05 | <0.001 |
| NS1 | 17.51 | 6.42 - 66.52 | <0.001 | 18.34 | 6.07 - 73.32 | <0.001 | 16.27 | 5.82 - 62.55 | <0.001 | 21.68 | 6.66 - 89.96 | <0.001 |
| Some RDT<sup>†</sup> | 5.71 | 3.31 - 9.81 | <0.001 | 2.67 | 1.34 - 5.44 | 0.0025 | 7.03 | 3.68 - 13.74 | <0.001 | 33.44 | 5.27 - 1374.7 | <0.001 |
| IgM OR IgG | 3.47 | 2.12 - 5.64 | <0.001 | 1.74 | 0.94 - 3.28 | 0.0146 | 4.09 | 2.33 - 7.17 | <0.001 | 11.83 | 3.47 - 61.93 | <0.001 |
| IgM OR NS1 | 5.01 | 3.12 - 8.02 | <0.001 | 3.12 | 1.68 - 5.85 | 0.0001 | 5.58 | 3.29 - 9.49 | <0.001 | 8.65 | 3.47 - 24.27 | <0.001 |
| IgG OR NS1 | 6.45 | 3.94 - 10.52 | <0.001 | 3.32 | 1.76 - 6.33 | 0.0001 | 7.87 | 4.44 - 14.02 | <0.001 | 14.45 | 4.79 - 57.79 | <0.001 |
| IgM (RTD OR ELISA) | 10.54 | 6.21 - 17.88 | <0.001 | 3.99 | 2.09 - 7.67 | <0.001 | 16.36 | 8.49 - 32.05 | <0.001 | 36.64 | 8.67 - 320.19 | <0.001 |

Dengue cases group is composed of cases classified as dengue without warning signs, dengue warning signs and severe dengue.
<sup>†</sup> Some RDT: IgM (Positive) OR RDT IgG (Positive) OR RDT NS1 (Positive)

**Table 4.** Multivariate analysis of risk factors associated with diagnosis in febrile patients

| Model | Multivariate analysis |  |  |  |  |  |  |  |  |  |  |  |
| --- | --- | --- | --- | --- | --- | --- | --- | --- | --- | --- | --- | --- |
|  | Dengue cases vs. OFI |  |  | Dengue vs.OFI |  |  | DWS vs. OFI |  |  | SD vs.OFI |  |  |
|  | aOR | IC 95% | *p* | aOR | IC 95% | *p* | aOR | IC 95% | *p* | aOR | IC 95% | *p* |
| **RDT IgM** | 2.63 | 1.59 - 4.33 | **<0.001** | 2.09 | 1.04 - 4.22 | 0.040 | 2.71 | 1.52 - 4.81 | 0.001 | 4.24 | 1.78 - 10.12 | 0.001 |
| Myalgia (Yes) | 1.87 | 1.04 - 3.38 | **0.038** | 2.21 | 0.95 - 5.15 | 0.067 | 1.94 | 0.97 - 3.89 | 0.062 | 1.50 | 0.54 - 4.16 | 0.434 |
| Abdominal pain (Yes) | 1.89 | 1.14 - 3.10 | **0.013** | 0.19 | 0.08 - 0.42 | **<0.001** | 4.46 | 2.48 - 8.03 | 0.000 | 2.93 | 1.24 - 6.92 | 0.014 |
| Platelets <140.000/mm<sup>3</sup> | 2.19 | 1.31 - 3.67 | **0.003** | 0.95 | 0.47 - 1.93 | 0.897 | 3.15 | 1.74 - 5.69 | 0.000 | 3.99 | 1.59 - 10.00 | 0.003 |
| **RDT NS1** | 18.09 | 5.52 - 59.34 | **<0.001** | 15.08 | 4.09 - 55.49 | **<0.001** | 17.99 | 5.18 - 62.52 | **<0.001** | 29.2 | 6.84 - 124.65 | **<0.001** |
| Myalgia (Yes) | 2.06 | 1.09 - 3.85 | **0.024** | 1.88 | 0.76 - 4.66 | 0.170 | 2.15 | 1.01 - 4.59 | **0.047** | 2.21 | 0.62 - 7.83 | 0.219 |
| Abdominal pain (Yes) | 2.00 | 1.19 - 3.37 | **0.009** | 0.21 | 0.09 - 0.49 | **<0.001** | 4.92 | 2.59 - 9.32 | **<0.001** | 3.70 | 1.34 - 10.21 | 0.011 |
| Platelets <140.000/mm<sup>3</sup> | 2.35 | 1.38 - 3.99 | **0.032** | 0.85 | 0.40 - 1.82 | 0.682 | 3.31 | 1.77 - 6.18 | **<0.001** | 2.70 | 0.99 - 7.39 | 0.052 |
| **RDT (IgM OR NS1)** | 4.22 | 2.49 - 7.13 | **<0.001** | 2.98 | 1.40 - 6.33 | 0.004 | 4.27 | 2.32 - 7.85 | **<0.001** | 12.05 | 3.79 - 38.34 | **<0.001** |
| Myalgia (Yes) | 1.84 | 1.00 - 3.39 | **0.049** | 2.15 | 0.91 - 5.08 | 0.082 | 2.01 | 0.98 - 4.12 | 0.058 | 1.47 | 0.51 - 4.23 | 0.474 |
| Abdominal pain (Yes) | 2.00 | 1.19 - 3.35 | **0.008** | 0.20 | 0.09 - 0.45 | **<0.001** | 4.65 | 2.53 - 8.55 | **<0.001** | 3.28 | 1.34 - 8.05 | 0.009 |
| Platelets <140.000/mm<sup>3</sup> | 1.89 | 1.11 - 3.23 | **0.020** | 0.84 | 0.41 - 1.75 | 0.650 | 2.83 | 1.54 - 5.21 | 0.001 | 3.75 | 1.43 - 9.83 | 0.007 |
| **IgM (RDT OR ELISA)** | 9.39 | 5.13 - 17.19 | **<0.001** | 3.98 | 1.81 - 8.75 | 0.001 | 12.22 | 5.86 - 25.49 | **<0.001** | 62.93 | 7.86 - 503.94 | **<0.001** |
| Myalgia (Yes) | 2.09 | 1.07 - 4.09 | 0.031 | 2.65 | 1.05 - 6.66 | 0.039 | 1.98 | 0.89 - 4.39 | 0.094 | 1.22 | 0.38 - 3.972 | 0.737 |
| Abdominal pain (Yes) | 1.77 | 1.00 - 3.12 | 0.048 | 0.19 | 0.08 - 0.45 | **<0.001** | 4.64 | 2.343 - 9.16 | **<0.001** | 4.20 | 1.51 - 11.72 | 0.006 |
| Platelets <140.000/mm<sup>3</sup> | 1.41 | 0.77 - 2.59 | 0.271 | 0.72 | 0.33 - 1.56 | 0.403 | 2.34 | 1.17 - 4.66 | 0.016 | 4.97 | 1.65 - 14.97 | 0.004 |

Dengue cases group is composed of cases classified as dengue without warning signs, dengue warning signs and severe dengue.

Therefore, using the results of the IgM RDT, NS1 RDT, and the combination of IgM RDT or NS1 RDT with the mentioned clinical variables allowed to differentiate between dengue cases and OFI cases (Table 4).

The significant variables from the regression analysis were used to establish decision trees with the RDT results representing the first branch, and the presence or absence of clinical signs the secondary branches. For example, for the IgM RDT decision tree, compared to the reference standard used to define dengue cases, we could detect 72.4% of the cases, while for the NS1 RDT decision tree, the percentage of detected dengue cases was 37%. The decision tree using the combined positive cases from the IgM RDT OR NS1 RDT and the combined positive cases from RDT NS1 or IgM ELISA, we could detect dengue cases in 81.6% and 90.6% of the cases, respectively (Fig 5). Contrarily, positive RDT IgG did not improve the detection of dengue cases.

**Figure 5.**
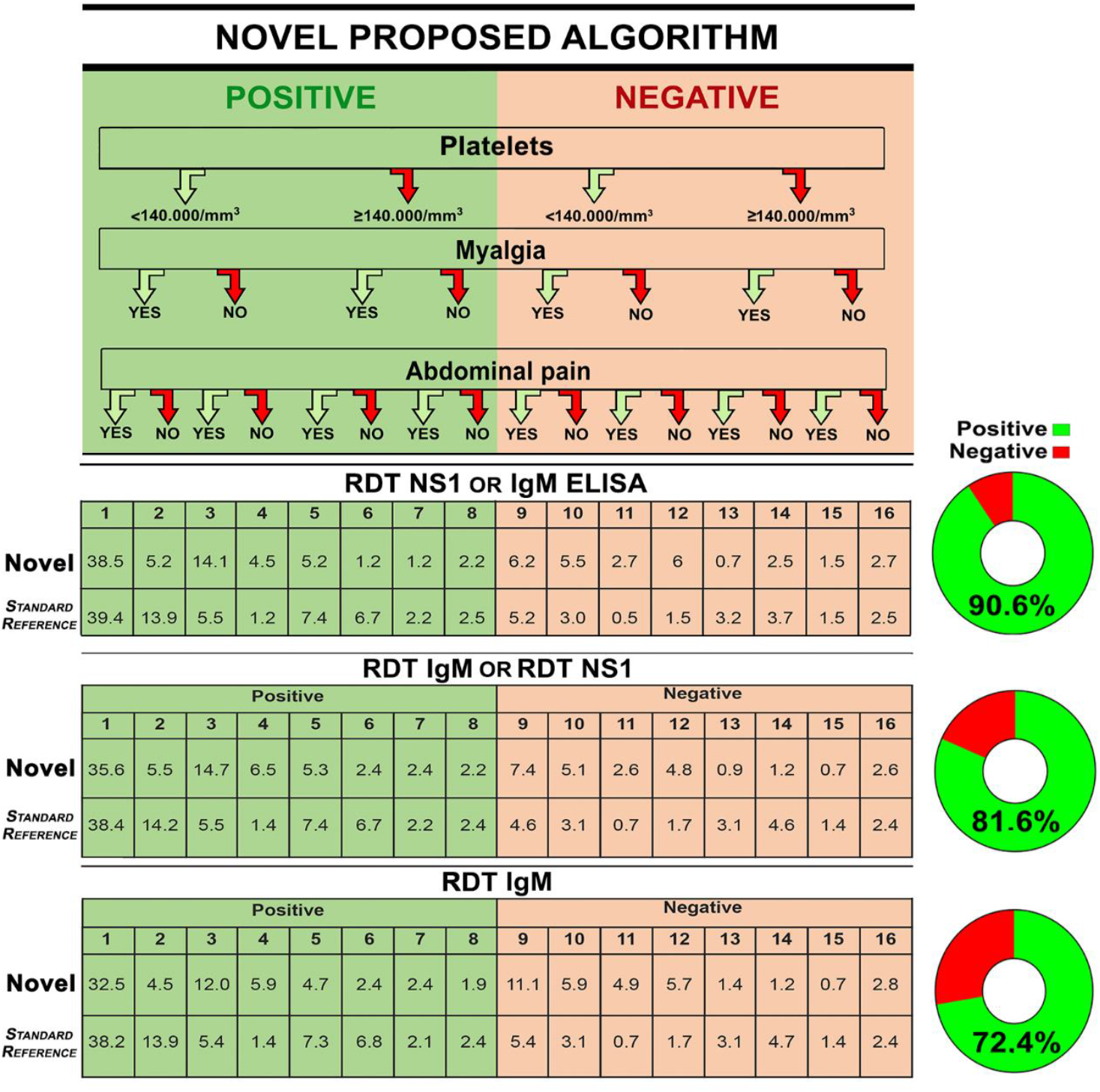
Decision tree for diagnosis of dengue cases using clinical and laboratory variables.

The area highlighted in green represents the dengue cases identified when applying the diagnostic algorithm with a positive test result (Columns 1 to 8). The area highlighted in pink represents the samples that present a negative test result and meet some of the clinical criteria proposed in the algorithm (Columns 9 to 16). The tables show the percentages of cases detected by the proposed algorithm, identified as Novel, (Upper row) and the percentages of dengue cases identified by standard reference (Lower row). The pie shows the percentage of cases detected by the new algorithm respect to the cases identified by the reference standard.

## Discussion

Based in a robust dengue case definition, in the present study, we challenge both serology and clinical characteristics to improve the case confirmation; a combined strategy yielded high sensitivity and specificity rates, that could help to clinicians in underdeveloped countries gain better opportunity and accuracy to confirm dengue cases in endemic areas.

The evaluation of diagnostic algorithms for dengue is challenging because of the dynamics of its clinical presentation [53, 58]. Similarly, hematological, biochemical, and immunological biomarkers assessed by clinicians in dengue patients show significant variations [73-75]. These variations are observed even in populations evaluated with similar characteristics such as age, time of disease, degree of severity, and genetic factors of individuals [54, 76, 77]. These differences make the inclusion of diagnostic algorithms in the care and management protocols for dengue cases challenging because their application creates a certain degree of uncertainty about their usefulness and effectiveness in the early identification of cases. However, the use of diagnostic algorithms may lead to a reduction in the percentage of individuals who develop complications and must bear the costs associated with the care of these patients.

In the present study, 61.7% of the population analyzed was under 15 years of age, which is consistent with the data reported during the same period by the Colombian Ministry of Health. During 2014-2015, the pediatric population registered the highest number of consultations for febrile syndrome in health institutions and the highest percentage of cases of dengue and severe dengue fever (40.8% and 34.2%, respectively) [44, 45].

Similar to the results reported by Tukasan et al. [78] we found differences in the temperature between SD cases and dengue cases (p=0.0008) as well as between DWS cases and dengue cases *(p=0*.*0051)*. Analysis of signs and symptoms such as headache, myalgia, nausea, and vomiting showed no significant differences between the dengue cases and OFI individuals [21]. These results are consistent with the research findings of Malhi et al. [79] and Chaloemwong et al. [25]. The clinical sign of abdominal pain on palpation in individuals with febrile syndrome allowed the classification of dengue cases according to the degree of severity and these were significantly different from OFI cases *(p=0*.*005)*. This symptom was not identified as a predictor variable for inclusion in the regression models and the design of the diagnostic algorithm. On the other hand, reported abdominal pain was found to be a predictor variable (OR: 2.36) and was considered in the development of the regression models. The study results are consistent with those reported by Da Silva et al., where the application of the WHO clinical criteria was found to be useful in identifying patients with warning signs and a high probability of developing severe dengue [80]. In the case of the laboratory diagnosis, the percentage of positive cases for dengue was significantly different between the IgM RDT (64.1%) and IgM ELISA (63.4%) tests (*p=0*.*033* and *p<0*.*0001*, respectively). The values obtained for sensitivity (70.5%) and specificity (56.9%) of the IgM rapid test, compared to the reference standard, were lower than the values obtained by other studies and ranged between 70 and 80% [63, 77]. These differences may be explained by variables such as the day of infection on which the diagnostic tests were performed, and the percentage of secondary infections identified; these two variables are critical in establishing this type of inference.

There are few studies where IgG Capture ELISA test has been included as a diagnostic marker and for confirmation of dengue cases [37, 38, 81, 82]. In the present study, we found that the frequency of positive samples in the IgG RDT was higher than that obtained in the IgG Capture ELISA test (66.1% and 52.4%). Further, the sensitivity and specificity of the IgG RDT were higher (72.6% and 55.2%) compared to IgG Capture ELISA (59.7% and 71.7%). However, these tests were not significant as predictor variables and therefore were not included in the regression models.

The percentage of positive results for IgM and IgG tests observed in our study differed from those reported in other studies; Senaratne et al. found that the percentage of samples positive for IgM and IgG was 61.9% and 83.9%, respectively [83]. Upon comparing this study with ours, we observed the reason for this was the differences in three population variables, namely, age of the subjects, place of residence, and duration of residence in the place, between the two studies. Further, the percentage of individuals under 5 years of age included in this study (19.8%) indicated that there is a high probability that children diagnosed with dengue experience primary infections.

Detection of the NS1 protein in patient samples has become a popular tool in the early diagnosis of DENV infection. In this study, we found that the sensitivity of NS1 RDT (37.8%) and ELISA test for NS1 (39.3%) was similar to that reported by other studies from Colombia [46, 50] and Singapore [84].

However, our results differ from the findings of studies conducted in Vietnam [24, 60, 74] and Malaysia [85], where the sensitivity of these tests has been reported to be over 80%. These differences in the sensitivity may be related to factors such as the assay type (ELISA or RDT) [46, 47, 50] and the frequency of DENV-2 infections, since a loss of sensitivity of NS1 detection has been reported in previous studies when this serotype was involved [47, 49, 60, 86]. Additionally, a low sensitivity has been observed in secondary infections because of the presence of NS1 antibodies from a previous infection, which may block the antigen during the test processing [46, 84, 87].

In our analysis, we found that the combination of NS1 RDT with the IgM ELISA test along with clinical variables led to an increase in the predictive values (AUC 0.932) compared to the reference standard, along with a higher sensitivity (90.3%) and specificity (96.2%). These results are consistent with the results seen in a study by Clemen et al. in Colombia [49]. They observed an increase in the sensitivity (between 67.2% and 79.5%) when IgM RDT and NS1 were combined with the negative clinical diagnosis (no clinical criteria for dengue cases). In contrast, they found that combining positive clinical diagnosis with the RDT yielded a decrease in the sensitivity (less than 35%) and an increase in the specificity (from 66.3% to 98.7% and 97.3% respectively).

The regression models developed in our study included not only clinical variables but also combined diagnostic tests and hematological variables such as leukocyte and platelet count for the algorithm design. A similar analysis was performed in a Taiwanese study, which identified the usefulness of combining laboratory variables such as leukocyte and platelet count, liver function tests, and coagulation profile in laboratory-confirmed cases of dengue. They showed that the sensitivity was low (49.5%) with a high positive predictive value [75]. Similarly, the model evaluated by Diaz et al. found that the clinical manifestations of the disease were not predictive in a multivariate analysis, while some variables that occur less frequently in dengue patients (e.g., somnolence) were crucial as predictors for dengue diagnosis [57].

We found differences between the three diagnostic groups for sociodemographic variables, day of illness, and leukocyte count (p<0.05). However, these variables were not found to be significant predictors and were not included in the regression models. On the contrary, in a study by Tuan et al. in Vietnam [60], age, leukocyte count, and platelet count were predictor variables within the model for early dengue classification. A later study conducted by this that group to establish a predictive model for severe dengue found that NS1 alone is not helpful for the classification. Hence, the reported model included this test in addition to variables such as vomiting, platelet count, and aspartate aminotransferase (AST) levels [24].

In this study, all the diagnostic tests when applied individually identified a lower percentage of dengue cases compared to the reference standard (S1 Table). For this reason, the models analyzed in the present study included a combination of clinical signs, hematological variables, and diagnostic tests, applied as unique tests as well in combinations. Our results showed that two of the models includeding a combination of clinical and hematological variables, with two RDT (IgM and NS1) or the combination of NS1 RDT with one of the tests considered as the standard reference (IgM ELISA), yielded a higher rate of case confirmation. These findings contrast to the usual method used to study dengue cases in endemic areas, where only the IgM ELISA test is performed.

The first algorithm can be used in higher-level health care institutions that perform IgM ELISA testing while applying the NS1 RDT simultaneously. The second algorithm can be used in health institutions in endemic areas because it is a combination of accessible, low-cost diagnostic tests, which complement the clinical information obtained during the evaluation of a patient; this would allow early diagnosis for the management and treatment of the disease. The application of this algorithm would increase the number of confirmed cases since the combined use of the tests would be able to detect cases that IgM ELISA might have missed and were diagnosed as other febrile syndromes. Our results differ from data reported by Jaenisch et al. [56], in individuals over 5 years of age from eight countries in Asia and Latin America. Although the work done by these authors includes the analysis of clinical and laboratory components to establish risk factors [56, 76], the diagnostic algorithm of our study additionally includes results from different tests.

Interestingly, we observed that some of the signs and symptoms included in the WHO definition for dengue (e.g., myalgia, hepatomegaly) were observed in both dengue cases and individuals diagnosed with OFI. Therefore, our findings highlight the fact that clinical diagnosis alone does not confirm true dengue cases and needs to be complemented by laboratory diagnostics. We also demonstrate the usefulness of rapid tests used together with IgM ELISA to confirm a higher percentage of dengue cases.

Among the main strengths of our work is the complete clinical information collected from our patients during their care at the institution and the various laboratory diagnostic tests used to confirm cases. Additionally, the evaluation of patients and the design of the algorithm applied during the period before the circulation of other arboviruses minimizes the possible interaction of diagnostic confounding variables. However, the limitations of our work include a low volume of serum samples obtained during the febrile phase, especially in patients under five years of age, which limited the application of the full panel of diagnostic tests, that may have resulted in a loss of individuals in the multivariate analysis.

In conclusion, our findings show that clinical diagnosis alone is not useful to confirm dengue cases and needs to be complemented laboratory tests. Further, the algorithms demonstrate the usefulness of the rapid test in combination with IgM ELISA to confirm a higher percentage of dengue cases, and we suggest that IgM RDT or NS1 RDT with some clinical variables allowed differentiate between dengue and OFI cases.

Considering that 70% of the individuals in our study were from the pediatric population (under 15 years), we suggest the evaluation and application of our algorithms in a multicenter study in both pediatric and adult populations, to identify their usefulness in the diagnosis of dengue cases in endemic areas.

## Data Availability

All data is enclosed in the manuscript file

## Funding

The project was funded by the Vicerrectoría de Investigaciones, Universidad El Bosque (PCI2013-457, PCI 2017-9423, PCI 2018-10297, research fellowship program) and the Colombian Department for Science, Technology and Innovation (Minciencias) project: Red de Investigación Multidisciplinaria para la prevención y control de Enfermedades Transmitidas por Vectores Red ETV (Project 360-2011).

## Acknowledgments

We are grateful to Miguel Otero Cadena from Vicerrectoría de Investigaciones of Universidad El Bosque, and members from Grupo de Virología of Universidad El Bosque. We also thank to Caucaseco Research Center (CICC), health care professionals from Hospital Universitario de la Samaritana – Unidad Funcional Girardot, for their help in the phase of patient recruitment, follow up and sample collection. Finally, we are extremely grateful to the study participants adults, children and their parents.

## Competing interests

The authors have declared that no competing interests exist.

## Supporting information

**S1 Table. S1 Table**. Statistical measures of dengue diagnostic tests. (DOCX)

**S2 Table**. Multivariate analysis of clinical and laboratory variables associated with diagnosis in febrile patients. (DOCX)

**S1 Fig**. Serotypes of DENV identified from dengue samples. (DOCX)

## Author contributions

**Carolina Coronel-Ruiz** Patient recruitment, samples collection and processing. Data management, analysis, and interpretation. Manuscript writing

Affiliation: Universidad El Bosque. Vicerrectoría de Investigaciones, Grupo de Virología. Bogotá, Colombia

**Myriam L. Velandia Romero** Project manager, Data interpretation. Manuscript revision Affiliation: Universidad El Bosque. Vicerrectoría de Investigaciones, Grupo de Virología. Bogotá, Colombia

**Eliana Calvo:** Molecular tests processing. Data interpretation. Affiliation: Universidad El Bosque. Vicerrectoría de Investigaciones, Grupo de Virología. Bogotá, Colombia

**Sigrid Camacho-Ortega:** Samples collection. Serology testing and molecular biology processing.

Affiliation: Universidad El Bosque. Vicerrectoría de Investigaciones, Grupo de Virología. Bogotá, Colombia.

**Shirly Parra-Alvarez:** Samples collection and serology testing.

Affiliation: Universidad El Bosque. Vicerrectoría de Investigaciones, Grupo de Virología. Bogotá, Colombia

**Edgar Beltrán-Zuñiga:** Molecular test processing.

Affiliation: Universidad El Bosque, Vicerrectoría de Investigaciones. Grupo de Virología, Bogotá, Colombia.

**María Angélica Calderón-Peláez:** Molecular test processing.

Affiliation: Universidad El Bosque. Vicerrectoría de Investigaciones, Grupo de Virología. Bogotá, Colombia

**Fabián Cortés-Muñoz:** Data analysis and interpretation. Manuscript revision Affiliation: Universidad El Bosque. Vicerrectoría de Investigaciones. Bogotá, Colombia

**Juan Pablo Rojas Hernández:** Patients clinical diagnosis and case classification. Affiliation: Fundación Clínica Infantil Club Noel. Cali, Colombia

**Syrley Velasco Alvarez:** Patients collection, samples collection. Affiliation: Hospital Universitario de La Samaritana. Bogotá, Colombia **Alfredo Pinzón Junca:** Patients collection. Data interpretation.

Affiliation: Hospital Universitario de La Samaritana. Grupo Investigación en Riesgo Cardio-Vascular, Trombosis y Anticoagulación (RICAVTA), Bogotá, Colombia

**Jaime E. Castellanos:** Principal investigator, Data analysis and interpretation. Manuscript writing and revision. Affiliation: Universidad El Bosque. Vicerrectoría de Investigaciones, Grupo de Virología. Bogotá, Colombia

## Notes

### Competing Interest Statement

The authors have declared no competing interest.

### Funding Statement

The project was funded by the Vicerrectoria de Investigaciones, Universidad El Bosque (PCI2013-457, PCI 2017-9423, PCI 2018-10297, research fellowship program) and the Colombian Department for Science, Technology and Innovation (Minciencias) project: Red de Investigacion Multidisciplinaria para la prevencion y control de Enfermedades Transmitidas por Vectores Red ETV (Project 360-2011).

### Author Declarations

This study was revised and approved by the Ethical Committee of Hospital Universitario de La Samaritana, Bogota (Resolution 7 and 9 of 2013 and 2014 respectively).

