## Supplementary data for "Improving dengue case confirmation by combining rapid diagnostic test, clinical, and laboratory variables"

#### Supporting information

**S1 Table.** Statistical measures of dengue diagnostic tests

| Variable | Sensitivity (%) | Sensitivity 95% CI | Specificity (%) | Specificity 95% CI | PPV (%) | PPV 95% CI | NPV (%) | NPV 95% CI | AUC |
|---|---|---|---|---|---|---|---|---|---|
| RDT IgM | 70.5 | 65.7 - 74.8 | 56.9 | 47.8 - 65.5 | 84.4 | 80.0 - 87.9 | 36.9 | 30.1 - 44.1 | 0.637 |
| IgM ELISA* | 81.6 | 77.3 - 85.2 | 100 | 96.5 - 100 | 100 | 98.8 - 100 | 60.8 | 53.4 - 67.7 | 0.908 |
| RDT IgG | 72.6 | 67.9 - 76.8 | 55.2 | 46.1 - 63.9 | 84.2 | 79.9 - 87.8 | 37.9 | 30.9 - 45.4 | 0.639 |
| IgG Capture ELISA | 59.7 | 54.7 - 64.5 | 71.7 | 62.8 - 79.2 | 87.5 | 83.0 - 91.0 | 34.8 | 28.9 - 41.1 | 0.657 |
| RDT NS1 | 38.7 | 33.9 - 43.7 | 96.5 | 91.4 - 98.6 | 97.4 | 93.4 - 99.0 | 32.3 | 27.5 - 37.4 | 0.676 |
| NS1 ELISA | 39.3 | 34.0 - 45.0 | 96.6 | 82.8 - 99.4 | 99.2 | 95.4 - 99.9 | 13.3 | 9.4 - 18.6 | 0.679 |
| RT - PCR* | 58.1 | 53.1 - 63.0 | 100 | 96.8 - 100 | 100 | 98.3 - 100 | 42.4 | 36.8 - 48.3 | 0.791 |

Statistics measures of single tests applied
* Test were considered in the gold standard definition

| Variable | | Sensitivity (%) | Sensitivity 95% CI | Specificity (%) | Specificity 95% CI | PPV (%) | PPV 95% CI | NPV (%) | NPV 95% CI | AUC |
|---|---|---|---|---|---|---|---|---|---|---|
| **IgM** | RDT OR ELISA | 88.2 | 84.5 - 91.1 | 58.5 | 49.0 - 67.4 | 88.2 | 84.5 - 91.1 | 58.5 | 49.0 - 67.4 | 0.734 |
| **IgG** | RDT OR ELISA | 78.5 | 74.0 - 82.3 | 46.8 | 37.8 - 56.1 | 83.3 | 79.1 - 86.9 | 39.1 | 31.2 - 47.6 | 0.627 |
| **NS1** | RDT OR ELISA | 43.1 | 37.7 - 48.8 | 96.4 | 82.3 - 99.4 | 99.2 | 95.8 - 99.9 | 13.7 | 9.6 - 19.2 | 0.697 |

Positivity of each RDT OR the respective ELISA test

| Variable | | Sensitivity (%) | Sensitivity 95% CI | Specificity (%) | Specificity 95% CI | PPV (%) | PPV 95% CI | NPV (%) | NPV 95% CI | AUC |
|---|---|---|---|---|---|---|---|---|---|---|
| **ELISA** | IgM OR IgG Capture | 84.8 | 80.8 - 88.1 | 70.8 | 61.5 - 78.6 | 91.1 | 87.7 - 93.7 | 56.8 | 48.3 - 65.0 | 0.778 |
| | IgM OR NS1 | 97.0 | 94.4 - 98.4 | 82.1 | 64.4 - 92.1 | 98.3 | 96.1 - 99.3 | 71.9 | 54.6 - 84.4 | 0.896 |
| | IgG Capture OR NS1 | 85.7 | 81.2 - 89.2 | 24.1 | 12.2 – 42.1 | 92.1 | 88.4 - 94.7 | 14.0 | 7.0 - 26.2 | 0.549 |

Positivity of some of ELISA test combined applied

| Variable | Sensitivity (%) | Sensitivity 95% CI | Specificity (%) | Specificity 95% CI | PPV (%) | PPV 95% CI | NPV (%) | NPV 95% CI | AUC |
|---|---|---|---|---|---|---|---|---|---|
| IgM ELISA OR RT-PCR[Ɉ] | 99.2 | 97.6 - 99.7 | 100 | 96.5 - 100 | 100 | 99.0 - 100 | 97.2 | 92.2 – 99.1 | 0.996 |
| IgG Capture OR RT-PCR[Ɉ] | 87.7 | 84.0 - 90.7 | 70.5 | 61.5 - 78.2 | 90.9 | 87.4 - 93.4 | 63.2 | 54.5 - 71.1 | 0.791 |
| NS1 OR RT-PCR[Ɉ] | 64.8 | 59.2 - 70.0 | 96.6 | 82.8 - 99.4 | 99.5 | 97.1 - 99.9 | 21.1 | 15.0 - 28.7 | 0.807 |

[Ɉ]Combination of RT-PCR positivity OR ELISA test applied positivity

| Variable | Sensitivity (%) | Sensitivity 95% CI | Specificity (%) | Specificity 95% CI | PPV (%) | PPV 95% CI | NPV (%) | NPV 95% CI | AUC |
|---|---|---|---|---|---|---|---|---|---|
| RDT IgM | 70.5 | 65.7 - 74.8 | 56.9 | 47.8 - 65.5 | 84.4 | 80.0 - 87.9 | 36.9 | 30.1 - 44.1 | 0.637 |
| IgM (RDT OR ELISA) | 88.2 | 84.5 - 91.1 | 58.5 | 49.0 - 67.4 | 88.2 | 84.5 - 91.1 | 58.5 | 49.0 - 67.4 | 0.734 |
| RDT IgM OR IgG Capture ELISA | 83.7 | 79.7 - 87.1 | 41.4 | 32.7 - 50.7 | 82.8 | 78.7 - 86.3 | 43.0 | 34.0 - 52.5 | 0.626 |
| RDT IgM OR NS1 ELISA | 83.3 | 78.7 – 87.1 | 55.2 | 37.5 - 71.6 | 95.1 | 91.7- 97.1 | 24.2 | 15.15 – 35.8 | 0.692 |
| RDT IgM OR RT-PCR | 90.0 | 86.6 - 92.6 | 57.0 | 47.8 - 65.7 | 87.5 | 83.8 - 90.4 | 63.1 | 53.5 - 71.8 | 0.735 |
| RDT (IgM OR IgG) | 84.1 | 80.1 - 87.4 | 39.7 | 31.2 - 48.8 | 82.1 | 78.0 - 85.6 | 43.0 | 34.0 - 52.5 | 0.619 |
| RDT (IgM OR NS1) | 80.5 | 76.2 - 84.2 | 54.8 | 45.7 - 63.6 | 85.5 | 81.4 - 88.7 | 46.0 | 37.9 - 54.3 | 0.677 |

Single IgM positivity or combined with other RDT OR ELISA OR RT-PCR

| Variable | Sensitivity (%) | Sensitivity 95% CI | Specificity | Specificity 95% CI | PPV (%) | PPV 95% CI | NPV (%) | NPV 95% CI | AUC |
|---|---|---|---|---|---|---|---|---|---|
| RDT IgG | 72.6 | 67.9 - 76.8 | 55.2 | 46.1 - 63.9 | 84.2 | 79.9 - 87.8 | 37.9 | 30.9 - 45.4 | 0.639 |
| RDT IgG OR IgM ELISA | 90.1 | 86.6 - 92.7 | 55.7 | 46.2 - 64.8 | 87.7 | 84.1 - 90.6 | 61.5 | 51.5 – 70.6 | 0.729 |
| RDT IgG OR IgG Capture ELISA | 78.5 | 74.0 - 82.3 | 46.8 | 37.8 - 56.1 | 83.3 | 79.1 - 86.9 | 39.1 | 31.2 - 47.6 | 0.627 |
| RDT IgG OR NS1 ELISA | 88.3 | 84.2 - 91.5 | 27.6 | 14.7 - 45.7 | 92.7 | 89.0 – 95.1 | 18.6 | 9.7 - 32.6 | 0.580 |
| RDT IgG OR RT- PCR | 94.5 | 91.7 - 96.4 | 55.3 | 46.1 - 64.1 | 87.6 | 84.0 - 90.4 | 75.0 | 64.8 - 83.0 | 0.749 |
| RDT (IgG OR IgM) | 84.1 | 80.1 - 87.4 | 39.7 | 31.2 - 48.8 | 82.1 | 78.0 - 85.6 | 43.0 | 34.0 - 52.5 | 0.619 |
| RDT (IgG OR NS1) | 85.5 | 81.6 - 88.7 | 52.2 | 43.1 - 61.1 | 85.5 | 81.6 - 88.7 | 52.2 | 43.1 - 61.1 | 0.689 |

Single IgG positivity or combined with other RDT OR ELISA OR RT-PCR

### Improving dengue case confirmation by combining rapid diagnostic test, clinical, and laboratory variables

| Variable | Sensitivity (%) | Sensitivity 95% CI | Specificity (%) | Specificity 95% CI | PPV (%) | PPV 95% CI | NPV (%) | NPV 95% CI | AUC |
|---|---|---|---|---|---|---|---|---|---|
| RDT NS1 | 38.7 | 33.9 - 43.7 | 96.5 | 91.4 - 98.6 | 97.4 | 93.4 - 99.0 | 32.3 | 27.5- 37.4 | 0.676 |
| RDT NS1 OR IgM ELISA | 90.3 | 86.9 - 92.9 | 96.2 | 90.6 - 98.5 | 98.8 | 97.0 - 99.5 | 73.7 | 65.8 - 80.4 | 0.932 |
| RDT NS1 OR IgG Capture ELISA | 79.4 | 75.0 – 83.2 | 70.0 | 60.9 – 77.8 | 90.0 | 86.3 – 92.8 | 50.0 | 42.2 - 57.8 | 0.747 |
| RDT NS1 OR NS1 ELISA | 43.1 | 37.7 - 48.8 | 96.4 | 82.3 - 99.4 | 99.2 | 95.8 - 99.9 | 13.7 | 9.6 - 19.2 | 0.697 |
| RDT NS1 OR RT-PCR | 69.0 | 64.1 - 73.4 | 96.5 | 91.3 - 98.6 | 98.5 | 96.2 - 99.4 | 48.2 | 41.8 - 54.7 | 0.827 |
| RDT (NS1 OR IgM) | 80.5 | 76.2 - 84.2 | 54.8 | 45.7 - 63.6 | 85.5 | 81.4 - 88.7 | 46.0 | 37.9 - 54.3 | 0.677 |
| RDT (NS1 OR IgG) | 85.5 | 81.6 - 88.7 | 52.2 | 43.1 - 61.1 | 85.5 | 81.6 - 88.7 | 52.2 | 43.1 - 61.1 | 0.689 |

Single NS1 positivity or combined with other RDT OR ELISA OR RT-PCR

| Variable | Sensitivity (%) | Sensitivity 95% CI | Specificity (%) | Specificity 95% CI | PPV (%) | PPV 95% CI | NPV (%) | NPV 95% CI | AUC |
|---|---|---|---|---|---|---|---|---|---|
| Some RDT ǂ | 90.5 | 87.2 - 93.1 | 37.4 | 29.1 – 46.5 | 82.7 | 78.8 – 86.0 | 54.4 | 43.5 – 65.0 | 0.639 |
| Some RDT ǂ OR IgM ELISA | 95.4 | 92.8 - 97.1 | 38.1 | 29.4 - 47.6 | 84.5 | 80.8 - 87.7 | 70.2 | 57.3 - 80.5 | 0.668 |
| Some RDT ǂ OR IgG Capture ELISA | 93.6 | 90.7 - 95.7 | 30.9 | 23.0 - 40.1 | 82.2 | 78.3 - 85.5 | 58.6 | 45.8 - 70.4 | 0.622 |
| Some RDT ǂ OR NS1 ELISA | 94.0 | 90.7 - 96.2 | 21.4 | 10.2 - 39.5 | 92.7 | 89.3 - 95.2 | 25.0 | 12.0 - 44.9 | 0.577 |
| Some RDT ǂ OR RT-PCR | 97.4 | 95.2 - 98.6 | 36.3 | 28.0 - 45.5 | 83.6 | 79.8 - 86.8 | 80.4 | 67.5 - 89.0 | 0.668 |
| Two RDT ǂǂ (Positive) | 69.2 | 64.4 - 73.6 | 71.3 | 62.5 - 78.8 | 88.9 | 84.8 - 92.0 | 41.2 | 34.6 - 48.1 | 0.703 |
| All RDT ǂǂǂ (Positive) | 22.1 | 18.2 - 26.5 | 99.1 | 95.2 - 99.8 | 98.8 | 93.6 - 99.8 | 27.8 | 23.7 – 32.3 | 0.606 |

ǂ RDT IgM (Positive) OR RDT IgG (Positive) OR RDT NS1 (Positive)

ǂǂ (RDT IgM (Positive) AND RDT IgG (Positive) OR (RDT IgM (Positive) AND RDT NS1 (Positive)) OR (RDT IgG (Positive) AND RDT NS1 (Positive))

ǂǂǂ RDT IgM (Positive) AND RDT IgG (Positive) AND RDT NS1 (Positive)

### Improving dengue case confirmation by combining rapid diagnostic test, clinical, and laboratory variables

**S2 Table.** Multivariate analysis of clinical and laboratory variables associated with diagnosis in febrile patients.

| Model | Multivariate analysis | | | | | | | | | | | |
|---|---|---|---|---|---|---|---|---|---|---|---|---|
| | Dengue cases vs. OFI | | | Dengue vs.OFI | | | DWS vs. OFI | | | SD vs.OFI | | |
| | aOR | IC 95% | *p* | aOR | IC 95% | *p* | aOR | IC 95% | *p* | aOR | IC 95% | *p* |
| **RDT IgG** | 2.79 | 1.69 - 4.62 | <0.001 | 1.53 | 0.77 - 3.07 | 0.221 | 3.72 | 2.07 - 6.69 | <0.001 | 5.616 | 2.24 - 14.08 | <0.001 |
| Myalgia (Yes) | 1.61 | 0.89 - 2.89 | 0.116 | 2.14 | 0.91 - 4.99 | 0.080 | 1.56 | 0.77 - 3.16 | 0.219 | 0.983 | 0.35 - 2.78 | 0.973 |
| Abdominal pain (Yes) | 1.75 | 1.06 - 2.88 | 0.029 | 0.18 | 0.08 - 0.39 | <0.001 | 4.42 | 2.43 - 8.03 | <0.001 | 2.626 | 1.09 - 6.29 | 0.030 |
| Platelets <140.000/mm$^3$ | 2.20 | 1.32 - 3.69 | 0.003 | 1.04 | 0.52 - 2.07 | 0.922 | 3.18 | 1.74 - 5.79 | <0.001 | 4.707 | 1.85 - 12.00 | 0.001 |
| **Some RDT** | 4.24 | 2.37 - 7.60 | <0.001 | 2.16 | 0.98 - 4.76 | 0.057 | 4.99 | 2.46 - 10.17 | <0.001 | 28.31 | 3.61 - 221.89 | 0.001 |
| Myalgia (Yes) | 1.69 | 0.93 - 3.09 | 0.084 | 2.14 | 0.91 - 5.02 | 0.081 | 1.75 | 0.86 - 3.56 | 0.122 | 1.29 | 0.46 - 3.66 | 0.630 |
| Abdominal pain (Yes) | 1.79 | 1.07 - 2.97 | 0.026 | 0.19 | 0.08 - 0.42 | <0.001 | 4.09 | 2.24 - 7.44 | <0.001 | 2.66 | 1.09 - 6.47 | 0.031 |
| Platelets <140.000/mm$^3$ | 2.20 | 1.31 - 3.71 | 0.003 | 1.01 | 0.50 - 2.03 | 0.979 | 3.35 | 1.84 - 6.12 | <0.001 | 4.92 | 1.92 - 12.58 | 0.001 |
| **RDT (IgM OR IgG)** | 2.88 | 1.67 - 4.95 | <0.001 | 1.79 | 0.86 - 3.77 | 0.120 | 3.18 | 1.67 - 6.05 | <0.001 | 9.30 | 2.56 - 33.79 | 0.001 |
| Myalgia (Yes) | 1.72 | 0.96 - 3.10 | 0.070 | 2.16 | 0.93 - 5.03 | 0.074 | 1.73 | 0.87 - 3.46 | 0.120 | 1.35 | 0.49 - 3.74 | 0.566 |
| Abdominal pain (Yes) | 1.83 | 1.11 - 3.02 | 0.017 | 0.18 | 0.08 - 0.40 | <0.001 | 4.25 | 2.36 - 7.66 | <0.001 | 2.65 | 1.11 - 6.33 | 0.029 |
| Platelets <140.000/mm$^3$ | 2.28 | 1.37 - 3.81 | 0.002 | 1.05 | 0.53 - 2.09 | 0.893 | 3.38 | 1.87 - 6.09 | <0.001 | 4.72 | 1.87 - 11.94 | 0.001 |
| **RDT (IgG OR NS1)** | 4.95 | 2.91 - 8.42 | <0.001 | 2.57 | 1.23 - 5.38 | 0.012 | 5.86 | 3.11 - 11.03 | <0.001 | 15.19 | 4.23 - 54.50 | <0.001 |
| Myalgia (Yes) | 1.67 | 0.90 - 3.09 | 0.103 | 2.04 | 0.86 - 4.84 | 0.108 | 1.68 | 0.81 - 3.49 | 0.165 | 1.13 | 0.39 - 3.33 | 0.820 |
| Abdominal pain (Yes) | 1.75 | 1.04 - 2.93 | 0.035 | 0.19 | 0.86 - 4.84 | 0.000 | 4.19 | 2.27 - 7.77 | <0.001 | 2.69 | 1.08 - 6.70 | 0.033 |
| Platelets <140.000/mm$^3$ | 1.96 | 1.147 - 3.36 | 0.014 | 0.92 | 0.86 - 4.84 | 0.817 | 3.03 | 1.63 - 5.62 | <0.001 | 4.44 | 1.71 - 11.58 | 0.002 |

Dengue cases group is composed of cases classified as dengue without warning signs, dengue warning signs and severe dengue.

**Improving dengue case confirmation by combining rapid diagnostic test, clinical, and laboratory variables**

**S1 Fig.** Serotypes of DENV identified from dengue samples.

A

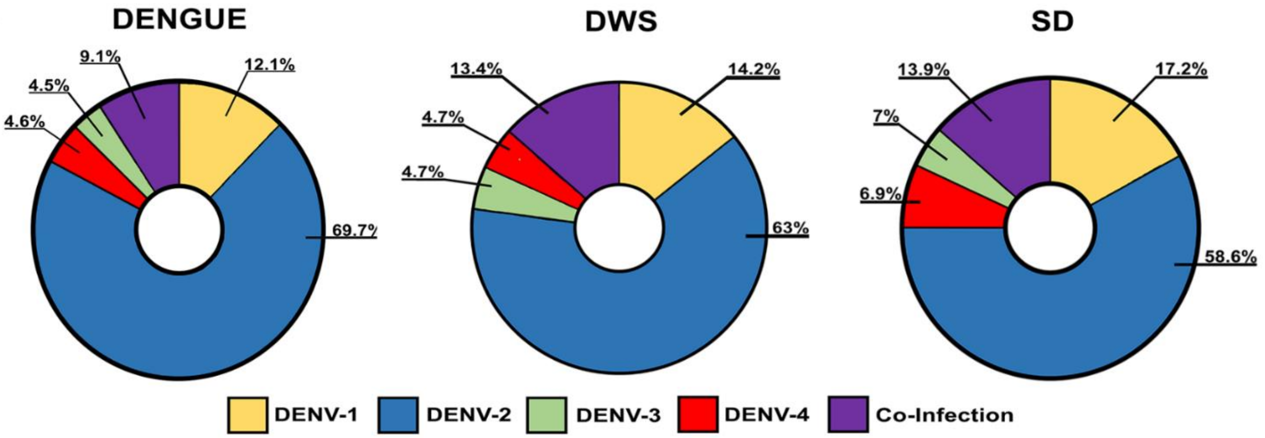


B

| Coinfection | Dengue | DWS | SD |
|---|---|---|---|
| DENV-1 **AND** DENV-2 | 3 (50.0%) | 7 (41.2%) | 2 (50.0%) |
| DENV-1 **AND** DENV-3 | 0 (0%) | 2 (11.8%) | 1 (25.0%) |
| DENV-1 **AND** DENV-4 | 1 (16.7%) | 0 (0%) | 0 (0%) |
| DENV-2 **AND** DENV-3 | 2 (33.3%) | 7 (41.2%) | 1 (25.0%) |
| DENV-3 **AND** DENV-4 | 0 (0%) | 1 (5.9%) | 0 (0%) |

**A.** Distribution of serotypes od DENV identified according to disease severity. Co-infections. Coinfections are defined by the detection of two DENV serotypes in the patient's sample. **B.** Description of DENV coinfections.
